# Epitweetr: Early warning of public health threats using Twitter data

**DOI:** 10.1101/2021.03.25.21254342

**Authors:** Laura Espinosa, Ariana Wijermans, Francisco Orchard, Michael Höhle, Thomas Czernichow, Pietro Coletti, Lisa Hermans, Christel Faes, Esther Kissling, Thomas Mollet

**Author notes:** These authors contributed equally to this manuscript (Laura Espinosa, Ariana Wijermans).

## Abstract

**Background:** ECDC performs epidemic intelligence activities to systematically collate information from a variety of sources, including Twitter, to rapidly detect public health events. The lack of a freely available, customisable and automated early warning tool using Twitter data, prompted ECDC to develop epitweetr.

The specific objectives are to assess the performance of the geolocation and signal detection algorithms used by epitweetr and to assess the performance of epitweetr in comparison with the manual monitoring of Twitter for early detection of public health threats.

**Methods:** Epitweetr collects, geolocates and aggregates tweets to generate signals and email alerts. Firstly, we evaluated manually the tweet geolocation characteristics of 1,200 tweets, and assessed its accuracy in extracting the correct location and its performance in detecting tweets with available information on the tweet geolocation. Secondly, we evaluated signals generated by epitweetr between 19 October and 30 November 2020 and we calculated the positive predictive value (PPV). Then, we evaluated the sensitivity, specificity and timeliness of epitweetr in comparison with Twitter manual monitoring.

**Findings:** The epitweetr geolocation algorithm had an accuracy of 30.1% and 25.9% at national and subnational levels, respectively. General and specific PPV of the signal detection algorithm was 3.0% and 74.6%, respectively. Epitweetr and/or manual monitoring detected 570 signals and 454 events. Epitweetr had a sensitivity of 78.6% [75.2% - 82.0%] and PPV of 74.6% [70.5% - 78.6%]; and the manual monitoring had a sensitivity of 47.9% [43.8% - 52.0%] and PPV of 97.9% [95.8% - 99.9%]. The median validation time difference between sixteen common events detected by epitweetr and manual monitoring was −48.6 hours [(−102.8) - (−23.7) hours].

**Interpretation:** Epitweetr has shown to have sufficient performance as an early warning tool for public health threats using Twitter data. Having developed epitweetr as a free, open-source tool with several configurable settings and a strong automated component, it is expected to increase its usability and usefulness to public health experts.

**Funding:** Not applicable

**Research in context:** *Evidence before this study:* Previous reviews have shown how social media, including Twitter, have been used for public health purposes. Most recent studies, in relation to the COVID-19 pandemic, have shown the added value of early warning tools based on Twitter and other social media platforms. They also noted the lack of an open-source tool for real-time monitoring and surveillance.

*Added value of this study:* Epitweetr is a free, open-source and R-based early warning tool for automatic Twitter data monitoring that will support public health experts in rapidly detecting public health threats. The evaluation of epitweetr presented in this study shows the strengths of the tool which include having good performance, high degree of automation, being a near-real-time tool and being publicly available with various customisable settings. Furthermore, it shows which are the areas of improvement for the next versions of epitweetr.

*Implications of all the available evidence:* This tool can be further developed to include more automation and machine learning components to increase usability and information processing time by users.

## Introduction

The European Centre for Disease Prevention and Control (ECDC) is a European Union (EU) agency aiming at strengthening Europe’s defences against infectious diseases. The article 3 of the ECDC Founding Regulation, the Decision Number 1082/2013/EU on serious cross-border threats to health and the ECDC Strategy 2021-2027 have established the detection of public health threats as a core activity of ECDC.

ECDC performs epidemic intelligence activities to systematically collate information from a variety of sources, which is then validated and analysed. The aim of epidemic intelligence at ECDC is to rapidly detect and assess public health events, focusing on infectious diseases, to ensure the EU’s health security^1^. Currently, ECDC monitors social media as part of its epidemic intelligence activities, in particular Twitter and Facebook^2^. In the past years, around one third of the signals detected at ECDC through epidemic intelligence activities originated from social media^3,4^. These platforms are often updated by local, national, and international health authorities capturing signals from small areas where media coverage is insufficient.

There have been other attempts to use social media data for the automatic early detection of signals of public health threats^5–8^ and a review of the use of Twitter for public health surveillance was published in December 2018^9^. However, this extensive review mainly targeted the monitoring of already detected outbreaks through Twitter, without fully covering ongoing monitoring of social media for early detection of public health threats. In addition, the authors found out that the geolocation of tweets through geotagging remained a major challenge. Several other published studies have described the use of Twitter for outbreak investigation^10–12^ or for understanding the public perception of an epidemic^13,14^, but these did not provide insights on the possible use of social media for automated event detection and monitoring in real time.

In the context of the coronavirus disease (COVID-19) pandemic, social media have become a key tool for sharing and disseminating data. In 2021, a scoping review was carried out to examine studies related to COVID-19 and social media during the first year of the pandemic^15^. ‘Surveillance and monitoring’ was one of the six themes extracted from these studies and according to the authors, no real-time surveillance monitoring has been developed for COVID-19 using social media data. Likewise, Lopreite and colleagues^16^, retrospectively analysed Twitter data to uncover early warning signals of COVID-19 outbreaks in Europe in the winter season 2019-2020, showing the relevance and stressing the urgency of having these early warning systems in place to better identify public health threats that may proliferate almost undetected otherwise.

Noting the usefulness of having free, customisable and automated early warning tools using social media, in particular Twitter data, ECDC developed a prototype of an R-based tool in August 2018 for the early detection of public health threats using Twitter data. The prototype focussed on a Public Health Event of International Concern (PHEIC) that had a major attention in social media: Ebola virus disease. This prototype was further extended in October 2019 and January 2020 by the inclusion of two other PHEICs: poliomyelitis and COVID-19, respectively. After the favourable results of this prototype, ECDC developed a free, open-source tool named epitweetr to automatically monitor Twitter data for early warning of public health threats. The first version of this tool was published on the Comprehensive R Archive Network (CRAN) in October 2020^**Error! Bookmark not defined**.^.

The main objective of this study is to evaluate epitweetr, a new automatised, open-source, R-based tool for early detection of public health threats using Twitter data. The specific objectives are to assess the performance of the geolocation and signal detection algorithms used by epitweetr and to assess the performance of epitweetr in comparison with the manual monitoring of Twitter for early detection of public health threats.

## Methods

### epitweetr

Epitweetr^17^ collects Twitter data and metadata using the Twitter Standard Search API 1.1. It collects these data by sending queries according to a predetermined list of topics with related keywords. Throughout the time of this study period a list of 71 unique topics was used^18^.

In parallel with the Twitter data collection, epitweetr processes these data to geolocate tweets, aggregate them, detect signals and send these signals through email alerts (Figure 1)^19^.

**Figure 1.**
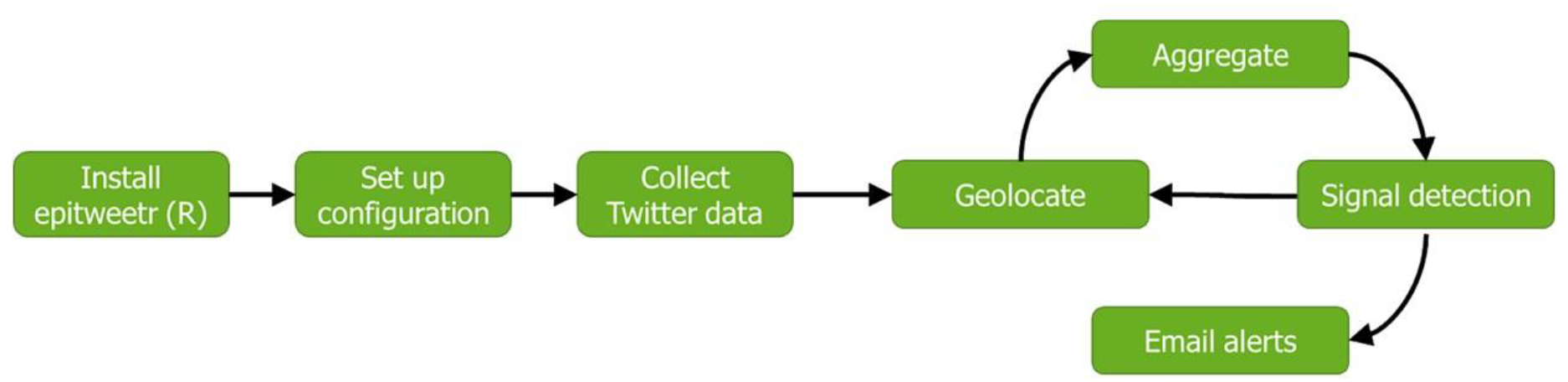
Epitweetr pipeline from installation to email alerts.

The geolocation process aims at collecting tweet and user location, with the tweet location being the primary location used for the signal detection. The tweet location is based on the location found in the tweet text or retweeted or quoted text. Epitweetr extracts the tweet location in two steps. In the first step, epitweetr looks for location candidates, splitting the tweet text into vectors and using language vectors from fasttext^18^ and a semi-supervised machine learning algorithm automatically trained with labelled datasets. In the second step, the text of these location candidates is matched against a reference database called geonames^18^ using a variant of vector space model (VSM) implemented on Lucene^20^. In each of these steps, a score is allocated and the location with the higher score is selected as candidate (first step) or as tweet geolocation (second step). For the user location, the location metadata available from the API is used. The best user location will be selected with a priority for user’s location at time of the tweet, followed by the self-declared user location or location as set in the public profile or the biography of the user.

The aggregation process creates the data shown in the three figures in the dashboard (Figure 2) based on Shiny web application framework^21^: time series of tweets per topic and location, map of tweet and/or user location, and the 20 most frequent words in the tweets per topic, time and location.

**Figure 2.**
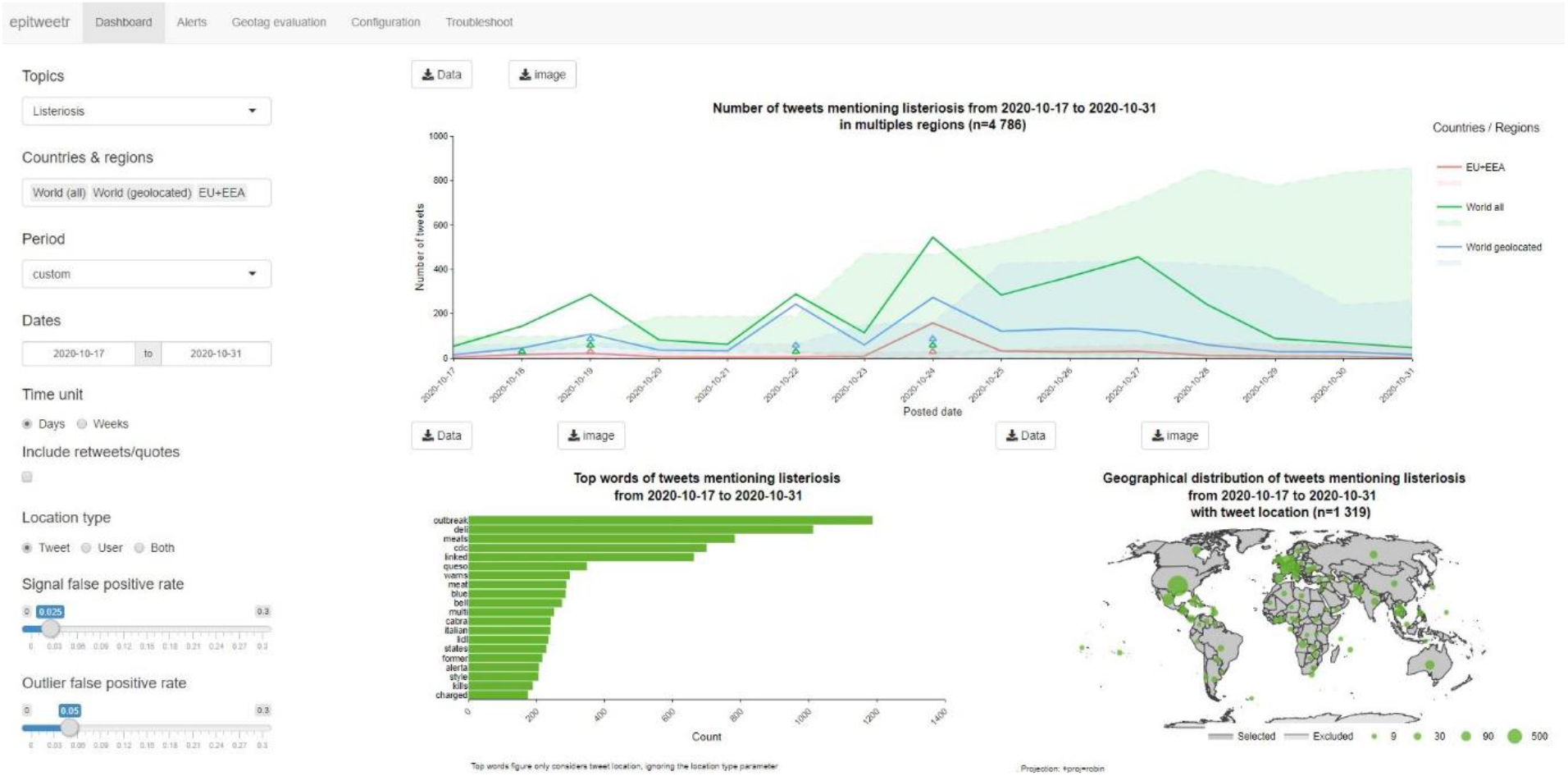
Epitweetr dashboard.

Epitweetr detects signals^19^, using the time series of tweets per topic and location (aggregated at country level). Each of the univariate time series is processed by a modified version of the Early Aberration Reporting System (EARS) algorithm^22^ based on linear modelling and as implemented in the R package ‘surveillance’^23^. Inspired by Farrington and colleagues^24^, the estimation of the threshold also downweights previous values if these are considered outliers. The algorithm then calculates a threshold for the expected tweet count for each topic and location (country level or higher) as a given quantile of the predictive distribution. If the threshold is exceeded a signal is created for that time series.

These signals are sent out in email alerts with the following variables available for each signal: date and hour of signal, geolocation of signal, most frequent words found in the tweet text, number of tweets, threshold, percentage of tweets from predetermined users and settings used at time of signal generation (Figure 3).

**Figure 3.**
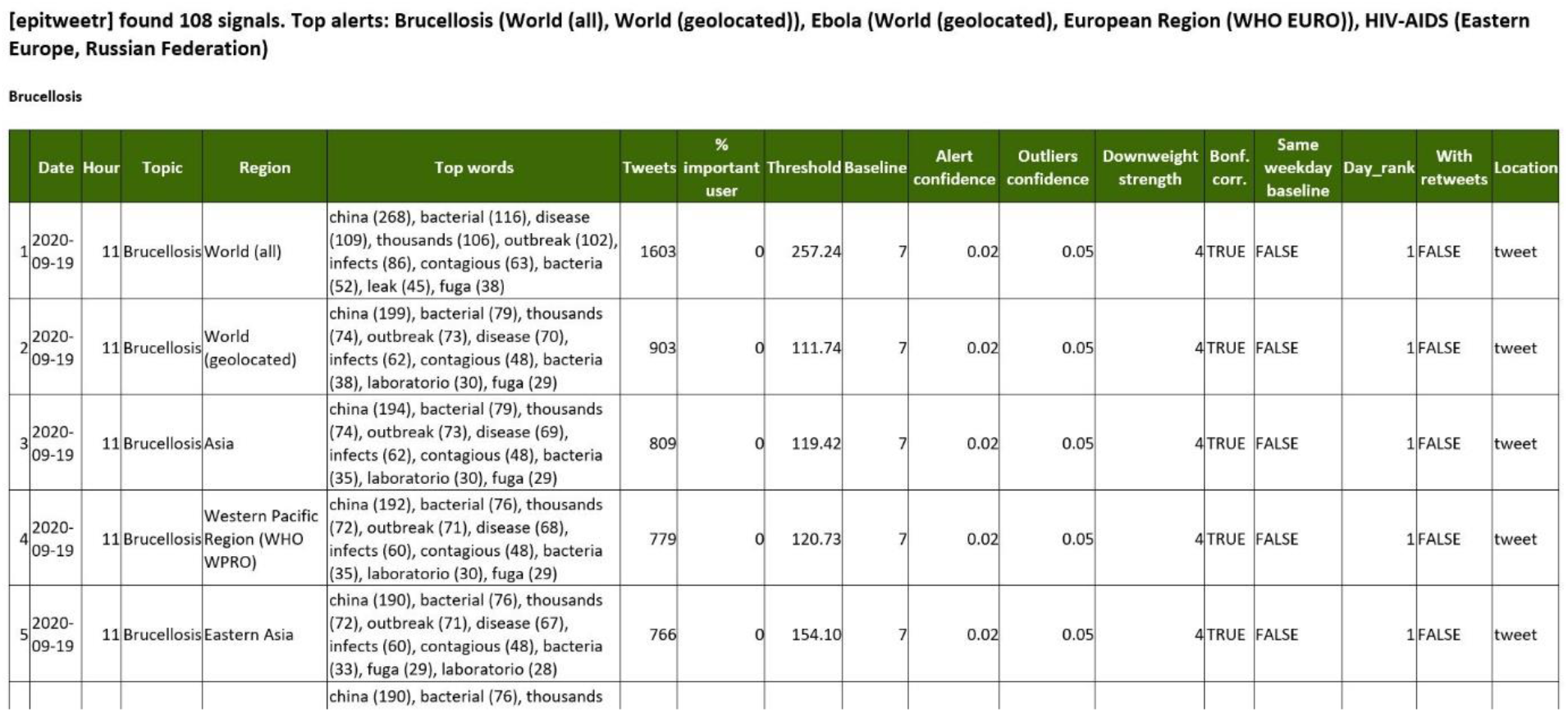
Example of email sent by epitweetr with detected signals.

### Evaluation of geolocation algorithm

We evaluated the primary geolocation of a random selection of 300 tweets per day collected on 11, 16, 19 and 22 September 2020, for a total of 1 200 tweets. All tweets were divided over two experts, with 150 tweets per day each. Each expert manually identified the best fitting geolocation at national and subnational level if such information was available.

The manual and epitweetr geolocations were then compared to evaluate the accuracy of the geolocation algorithm in extracting the correct location and the performance of the geolocation algorithm in deciding which tweets had available information to extract a location (hereafter referred as tweets with an extractable location) at national and subnational level. For each tweet, we defined a positive hit when a location could be extracted from the tweet and a negative hit when no location could be extracted from the tweet. For positive hits, tweets were considered true positives or false positives depending on whether epitweetr assigned a location for these tweets or not. For negative hits, tweets were considered true negatives or false negatives depending on whether epitweetr assigned a location for these tweets or not.

Using this information, we calculated the accuracy of extracting the correct tweet geolocation at national and subnational level. Furthermore, the following calculations were made regarding the performance in deciding which tweets had an extractable location at national and subnational level: sensitivity, specificity, positive predictive value (PPV), negative predictive value (NPV) and prevalence (number of tweets from which location could be extracted)^25^.

The average of the results from both individuals were calculated into an overall result. Additionally, these calculations were made according to the geolocation score dividing tweets in two groups: tweets with tweet geolocation score below and above 10.

Furthermore, we identified the most frequent errors made by the algorithm in extracting the correct location, and grouped them in two categories: tweets mentioning the US president and/or a ‘well established location’ (e.g. country names, country populations, US state names and capital cities). The same previously mentioned calculations were made to evaluate what would have been the performance of epitweetr geolocation algorithm if these locations had been extracted correctly by epitweetr.

### Evaluation of signal detection algorithm

We evaluated signals generated by epitweetr by topic, location and time during the working days between 19 October and 30 November 2020 to determine which were validated events. An event was an epitweetr signal that fulfilled ECDC criteria based on International Health Regulations and Decision 1082, and that was validated (i.e. deemed accurate and reliable information, and confirmed by or originated from an official source). The validation process can be automatic, when the information comes from an official source (e.g. Public Health Twitter accounts or international organisations websites), or manual using official sources, when the information comes from non-official sources or individual Twitter accounts.

Following the epidemic intelligence steps^26^, we further investigated signals that seemed to fulfil ECDC criteria according to the most frequent words, topic, location and number of tweets. We retrieved the events that could have triggered those signals and validated the information. On occasions, after retrieving the events the signals were discarded due to the additional information provided by the event itself that did not fulfil ECDC criteria.

We defined the general positive predictive value (*PPV*_*g*_) and specific PPV (*PPV*_*s*_) as:

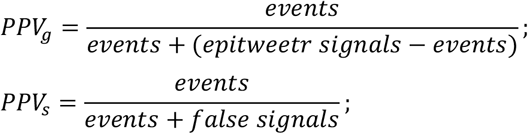

where *events* were epitweetr signals that fulfil ECDC criteria and were validated, *epitweetr signals* were the total number of signals detected by epitweetr during the study period, and *false signals* were evaluated signals that did not fulfil ECDC criteria and/or were not validated.

### Evaluation of epitweetr

We developed a study protocol to evaluate the sensitivity, specificity and timeliness of epitweetr in comparison to the manual monitoring of Twitter for early detection of public health threats (supplementary material).

### Ethics statement

Epitweetr collects Twitter data using the Twitter Standard Search API which provides relevant and only publicly available tweets matching a specific query from the previous seven days. These data are similar but not identical to the results provided by the Search User Interface feature in Twitter mobile or web clients.

## Results

### Evaluation of geolocation algorithm

At the country level, the epitweetr geolocation algorithm had an overall accuracy of 30.1%, while at the subnational level this was 25.9% (Table 1).

**Table 1.** Summarised results from the tweet geolocation evaluation.

|  |  | All results |  | >10 score |  |
| --- | --- | --- | --- | --- | --- |
| Level |  | Country | Subnational | Country | Subnational |
| Total tweets |  | 1200 | 1200 | 804 | 804 |
| Extracting correct location | Correct hits | 361 | 311 | 292 | 283 |
|  | Accuracy | 30.1% | 25.9% | 36.3% | 35.2% |
| Detecting tweets with extractable location | Sensitivity | 72.6% | 72.2% | 56.6% | 56.2% |
|  | Specificity | 51.6% | 50.6% | 69.2% | 68.3% |
|  | PPV | 74.9% | 74.2% | 75.7% | 74.9% |
|  | NPV | 48.6% | 48.1% | 48.6% | 48.1% |
|  | Prevalence | 66.6% | 66.3% | 62.8% | 62.7% |

If all tweets with the name of the US president had been correctly assigned (n=281 tweets), we observed an accuracy of 38.5%, a sensitivity of 76.0%, a specificity of 51.6% and a PPV of 75.8% at country level. If also all ‘well established locations’ had been correctly assigned (n=371), which includes besides the US president also country names, country populations, capitals, and US states, this would have led to an accuracy of 52.4%, a sensitivity of 88.8%, a specificity of 51.5% and a PPV of 78.5%.

### Evaluation of signal detection algorithm

During the study period, 11,313 signals were detected by epitweetr from which 448 were evaluated. From these evaluated signals, 334 were events and 114 were false signals.

From these 448 evaluated signals, 63 were related to COVID-19, including 48 events and 15 false signals. In addition, 49 of the 448 signals had only one tweet, including 24 events and 25 false signals.

Overall, the *PPV*_*g*_ was 3.0% and the *PPV*_*s*_ was 74.6%. The *PPV*_*s*_ for COVID-19 related events and for other events were 76.2% and 74.3%, respectively.

### Evaluation of epitweetr

In order to reach the minimum sample size, data were collected from 19 October to 30 November 2020. Overall, 570 signals were evaluated, including 122 signals detected by the manual method, 297 signals detected by epitweetr and 151 signals detected by both methods. From these 570 evaluated signals, 157 were related to COVID-19, including 120 signals detected by the manual method, 24 signals detected by epitweetr and 39 signals detected by both methods.

Overall, 454 events were detected, including 120 events detected by the manual method, 185 events detected by epitweetr and 149 events detected by both methods. From these 454 events, 157 were related to COVID-19, including 94 events detected by the manual method, 24 events detected by epitweetr and 39 events detected by both methods.

The number of signals and events, IRA and PPV of both methods are presented in Table 2.

**Table 2.** Summarised metrics of epitweetr evaluation.

| Variables | Manual method | epitweetr |
| --- | --- | --- |
| Number of signals | 188 | 448 |
| Number of events | 184 | 334 |
| IRA [95% CI] | 47.9% [43.8% - 52.0%] | 78.6% [75.2% - 82.0%] |
| PPV [95% CI] | 97.9% [95.8% - 99.9%] | 74.6% [70.5% - 78.6%] |

A total of 16 unique events were found by both methods, including 10 events related to COVID-19. The median of the validation time differences was −48.6 hours with an interquartile range (IQR) of −102.8 and −23.7 hours, showing an earlier validation of common events by epitweetr. Figure 4 shows the distribution of the validation time differences in hours.

**Figure 4.**
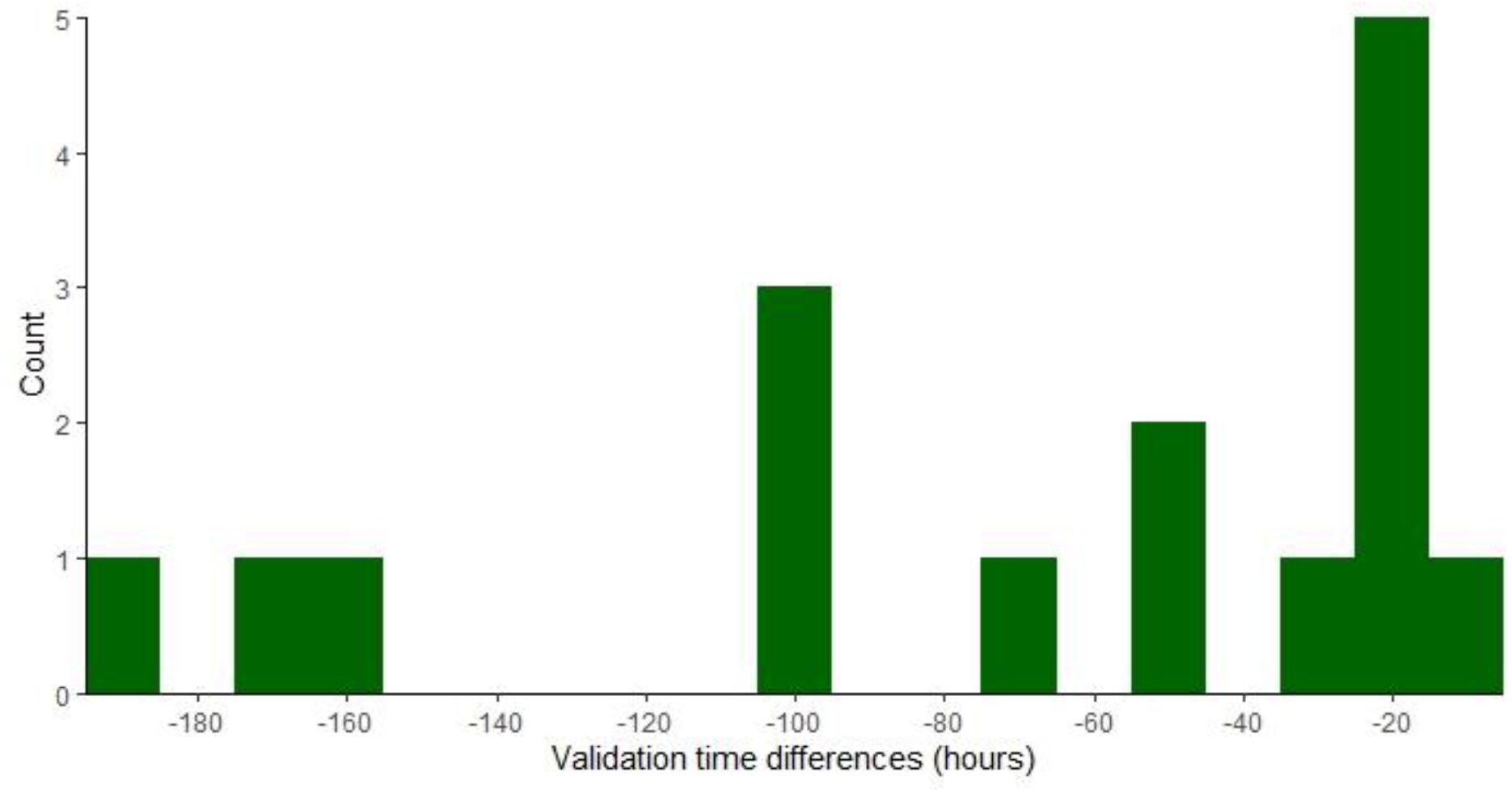
Histogram showing validation time differences in hours between epitweetr and manual method.

The Wilcoxon signed rank test showed that the validation time difference is significantly smaller than zero (p-value = 1.5 × 10^−5^), meaning that we validated events detected by epitweetr earlier than those detected by the manual method.

## Discussion

In this article we present the evaluation of epitweetr, a new automatised, open-source, R-based tool for early detection of public health threats using Twitter data. We developed this tool after finding a knowledge gap and after performing a feasibility study through a manual prototype of this type of tool.

Previous studies have shown the relevance of having an appropriate geolocation when using social media data to better understand where the event or threat is happening. We focus our geolocation evaluation on the tweet geolocation since it is more relevant for the objective of the tool (early detection of threats) providing a more accurate information related to the event rather than to the user. The tweet geolocation evaluation showed an accuracy of around 30%, being higher at national than at subnational level. The vast majority of the wrongly geolocated tweets were related to few recurrent errors from the algorithm such as US president and population citizens not being recognized (e.g. Trump, American, Chilean) or common words getting high priority (e.g. ‘real’ for the location Ciudad Real, Spain). By adding a supervised learning layer to the existing algorithm, the user could easily improve by training it and thus increasing substantially its accuracy and specificity. This was seen in the increased accuracy at national level from 30.1% to 38.5% and 52.4% respectively if the US president or a group of ‘well established locations’ could have been correctly identified. This naturally also increased the sensitivity (up to 88.8%). However, the PPV only increased slightly (from 74.9% to 75.8% and 78.5% respectively) as some of these tweets had already been assigned a location even if it was the wrong location.

Epitweetr users can modify the threshold used by the geolocation algorithm to prioritise sensitivity or accuracy and specificity. Our evaluation proved that using a score above 10 as threshold increased the accuracy from 30.1 to 36.3% and the specificity from 51.6% to 69.2%, but it also decreased the sensitivity as expected.

We decided to use the Early Aberration Reporting System (EARS) as a baseline for the signal detection algorithm since it is a well-established methodology initially developed by the United States Centers for Disease Control and Prevention (US CDC)^27^. The initial evaluation of the modified EARS algorithm stressed the need of having an adaptable system to adjust false positive rate and sensitivity. We addressed this need by adding configurable settings that can be adjusted even by topic. Additionally, we added a downweighting system for signal outliers, not to miss relevant signals afterwards. Likewise, we included Bonferroni correction with configurable settings since we are performing multiple tests at regional and country level^28^. Since the use of Bonferroni depends on the circumstances of the study, epitweetr users can activate or deactivate it according to their needs.

This modified EARS algorithm used by epitweetr for signal detection had a much higher sensitivity in comparison with the manual method and allowed to detect small signals containing only one tweet. In general, epitweetr detected more events than the manual method in a timelier manner.

When comparing the performance of epitweetr and the manual method for the COVID-19 signals and events, the latter detected more events. This can be explained by the fact that COVID-19 became a much-tweeted topic and the query used by epitweetr was too generic to detect these specific events. Creating more specific queries for COVID-19 (e.g. COVID-19 vaccines, COVID-19 treatment, COVID-19 schools) would have allowed to detect more events with epitweetr. This is relevant when a new event or threat is being monitored since a generic query can be used in the early stages and subqueries or more specific queries should be developed once the event becomes more popular and it is mentioned more frequently by Twitter users.

Epitweetr showed a lower positive predictive value in comparison with the manual method as expected for this signal detection algorithm. We developed epitweetr to detect small signals so having a very sensitive tool was a priority. There are some configurable settings that allow epitweetr users to modify the false positive rate of the tool. Furthermore, adding a machine learning module to epitweetr could increase the positive predictive value by combining supervised and unsupervised learning to continuously train the model and adapt to possible concept drifts in Twitter streams^29^ without jeopardising the sensitivity and IRA achieved by the underlying signal detection algorithm.

Throughout the evaluation, we found that having many configurable settings increased the flexibility of the tool and its ability to adapt to different contexts or specific uses. The dashboard of the epitweetr application is intended for testing all these settings before epitweetr users decide which values or parameters to use in their context. This decision will depend on the resources available, which relates to the specificity of the tool (e.g. experts available to assess all signals detected by epitweetr, including possible false signals) and the granularity required, which relates to the sensitivity of the tool (e.g. which would be the consequences of missing a small signal).

The main limitation of epitweetr relates to the variation in Twitter data dynamics and different scopes within early detection of threats that epitweetr users may have. We have overcome this by adding most of the parameters as configurable settings that can be changed not only for the tool itself but also depending on the topic. Likewise, the dashboard facilitates this decision showing the immediate results of choosing different parameters.

In conclusion, epitweetr has shown to have sufficient performance for early detection of public health threats using Twitter data. This type of tool has shown to be useful to public health experts. Moreover, publishing epitweetr in a public repository with several customisable settings allows other users to adapt the tool to their specific needs and, even, further develop this tool. Additionally, since epitweetr has a strong automated component providing outputs in a near-real-time manner, we believe it can become a useful tool in the daily public health practice of infectious disease event and threat detection.

## Data Availability

All code used by epitweetr are available as an R package from CRAN. Source maintenance and interaction occurs through the GitHub repository. The historical Twitter data used in the present analysis cannot be shared. However, a dataset with the anonymised signals and events detected by epitweetr and the manual method for these data is publicly available.

https://github.com/lauespinosa/epitweetr_evaluation

https://github.com/EU-ECDC/epitweetr

## Code and Data sharing

All code used by epitweetr are available as an R package from CRAN. Source maintenance and interaction occurs through the GitHub repository^18^. The historical Twitter data used in the present analysis cannot be shared. However, a dataset with the anonymised signals and events detected by epitweetr and the manual method for these data is publicly available^30^.

## Authors’ contributions

LE, AW and TM conceived epitweetr and developed its prototype. FO, EK, MH and TC developed epitweetr. PC, LH and CF developed the study protocol to assess the performance of epitweetr in comparison with the manual monitoring of Twitter data. LE and AW developed and performed the assessment of the geolocation and signal detection algorithms of epitweetr, and performed the assessment of epitweetr in comparison with the manual monitoring of Twitter data. LE and AW have verified the underlying data. LE and AW drafted the manuscript. All authors contributed to the interpretation of the data and editing of the final manuscript. All authors have seen and approved the manuscript. LE and AW contributed equally to this manuscript.

## Supplementary material

## Appendix A

### Additional methods: epitweetr evaluation

The manual monitoring (hereafter referred to as the manual method) consisted of screening twice a day most recent tweets posted by a list of over 100 validated Twitter users followed by ECDC EI team. In this method, we defined a signal as a tweet that fulfils ECDC criteria and required further action (e.g. validation of the information). The time of this tweet was recorded as the signal time an d, in case several Twitter accounts were tweeting about the same topic in the same screening round, the earliest set the signal time.

The epitweetr screening consisted of screening twice a day email alerts (i.e. unexpected increase in the number of tweets by topic, location and time) sent by epitweetr at approximately 4.30 Central European Time (CET) and 13.30 CET. In this method, we defined a signal as an alert for a specific topic, location and time which top words and other information included in the email suggested it fulfilled ECDC criteria. The time of the earliest alert of the day was recorded as the signal time.

In both epitweetr and the manual method, an event was a validated signal deemed trustful and reliable by an official source. The approximate time at which this validation was done was recorded as the event time, both in case of positive and negative validation.

Two EI experts screened during alternate weeks Twitter data twice a day using both methods and recorded signals and events detected by each method until the minimum sample size was achieved.

Considering there is no prior estimate available for the sensitivity and specificity and having a maximal marginal error of 0.15 for sensitivity and specificity, we defined the minimum sam ple size as:

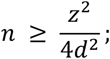

where *n* is the minimum sample size, *z* is the 97.5% percentile of the standard normal distribution, and *d* is the maximal marginal error.

In order to achieve a minimum sample size, a time period with the following criteria was selected: at least 43 different events found by any of the methods, at least 43 signals found by each method and at least 10 events found by both methods.

Since it is difficult to evaluate the classification accuracy of the generated events by the two methods, because no independent gold standard exists and there is no available information on all events that should be detected by both methods, we used instead an inter-rater agreement (IRA) between the two methods as a relative definition of sensitivity^1^. We defined the IRA of the manual method (*IRA*_*m*_) and the IRA of epitweetr (*IRA*_*e*_), with their 95% confidence interval (CI), as:

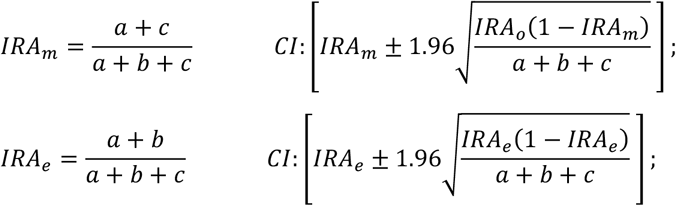

where *a* was the number of events detected by both methods, *b* was the number of events detected only by epitweetr, and *c* the number of events detected only by the manual method.

Since the estimation of the specificity was not feasible in this context, we calculated the PPV as the proportion of signals corresponding to a validated event. We defined the manual method PPV (*PPV*_*m*_) and the epitweetr PPV (*PPV*_*e*_), with their 95% CI, as:

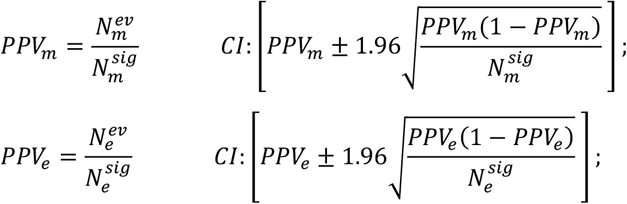

where 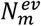 was the number of events detected by the manual method, 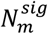 the number of signals detected by the manual method, 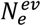 the number of events detected by epitweetr, and 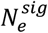 the number of signals detected by epitweetr.

We defined the timeliness as the difference between the validation time of events found by epitweetr and manual method. In case an event was detected in several days, only the earliest was kept for the analysis. We performed a descriptive analysis, including measures of central tendency and variability. Likewise, we performed a significance test using the signed rank test where the null hypothesis assumed there was no true difference and the alternative hypothesis assumed epitweetr had earlier validation times than the manual method.

